# Empowering Singaporeans to better manage chronic heart failure: Protocol for a mixed-methods study

**DOI:** 10.64898/2026.07.09.26357623

**Authors:** Bairavee, Yuxin Wang, Dharani Kanna Ravi, Audry Lee Shan Yin, Raymond Ching Chiew Wong, Seet Yoong Loh, Nicholas Graves, Sharon Sung, Sungwon Yoon, Derek J Hausenloy, Lian Leng Low, Khung-Keong Yeo, Kheng Leng David Sim, Yichi Zhang, Sanjeewa Kularatna, Sameera Senanayake

## Abstract

**Introduction:** The prevalence of chronic heart failure is increasing in Singapore and is associated with frequent hospitalisations, high costs, and impaired quality of life. Patient empowerment interventions for chronic diseases, which are structured approaches that enable patients to actively engage in and influence their care, have demonstrated promising effects on health-related outcomes. In chronic heart failure, however, many interventions focus on selected aspects of empowerment, and there remains limited synthesis of which approaches are most acceptable, preferred, and effective as comprehensive intervention packages. This protocol describes the methods for a study to identify an empowerment-based intervention for adults with chronic heart failure that is both contextually suitable and cost-effective in Singapore.

**Methods and analysis:** We will use a staged, sequential design comprising three objectives. Objective one is to conduct a systematic review and meta-analysis to synthesise international evidence of the effectiveness of empowerment-based interventions for adults with chronic heart failure. Objective two is to complete a mixed-methods study. Semi-structured interviews with patients and their caregivers, analysed using codebook thematic analysis, will identify empowerment-related needs, barriers and facilitators in local chronic heart failure care. Findings from the review and interviews will then inform a discrete choice experiment to elicit patients’ preferences for features of an empowerment-based intervention, analysed using multinomial logit and latent class models. Objective three is to conduct a cost-effectiveness analysis of the selected intervention compared with usual care, using a decision tree and Markov model from the perspective of the Singapore health system.

**Ethics and dissemination:** Ethics approval for the qualitative study has been obtained from the SingHealth Centralised Institutional Review Board (ECOS Ref: 2025-1152); ethics approval for the discrete choice experiment will be sought prior to recruitment. Findings will be disseminated through peer-reviewed publications and conference presentations.

**Registration details:** PROSPERO registration number: CRD420251249957; http://www.crd.york.ac.uk/PROSPERO

**ARTICLE SUMMARY:** *Strengths and limitations of this study:*

- The staged, sequential design integrates a systematic review, qualitative interviews, a discrete choice experiment and a cost-effectiveness analysis, with each component informing the next, combining international evidence with local stakeholder perspectives and patient preferences to inform a feasible, patient-centred intervention.
- Interviews with patients and caregivers across three institutions, offered in four languages and using purposive sampling by age, sex, language and socioeconomic status, aim to capture diverse local experiences of empowerment.
- Older adults and those with limited digital literacy may be under-represented or find the online discrete choice experiment difficult to complete, introducing potential selection bias and measurement error.
- Intervention uptake and adherence in the cost-effectiveness model will be based on stated rather than observed preferences, and effectiveness will be informed by existing evidence rather than a locally conducted trial.

## INTRODUCTION

Chronic heart failure (CHF) is a major global public health concern, associated with frequent hospitalisations, high healthcare costs, and reduced quality of life [1]. Its burden is expected to grow with population ageing, increasing multimorbidity, and lifestyle-related risk factors [1]. Singapore’s epidemiological data similarly reflect a substantial and growing CHF burden with clear implications for health system costs [2–4]. Sociocultural factors also shape the local context of CHF prevalence and management. These include weakening family caregiving capacity, documented health literacy gaps, including poor awareness of recommended lifestyle adjustments [5], and an ageing population.

Collectively, these epidemiological, economic, and social vulnerabilities highlight the need for approaches that actively support patients in managing their condition and sharing responsibility for care. In this context, patient empowerment interventions, which are structured approaches that enable patients to actively engage in and influence their care, have gained prominence in chronic disease management [6]. These interventions tend to improve clinical outcomes and quality of life, but empowerment remains conceptually heterogeneous and inconsistently defined across studies [6–7].

Specific to CHF, empowerment-based interventions typically include components such as education and cognitive-behavioural support [8–9]. Evidence suggests potential benefits for self-care behaviours, quality of life, and hospitalisation outcomes [8–9]. However, empowerment approaches often focus on selected components such as education or self-monitoring, though evidence suggests that more integrated, multi-disciplinary approaches may be more effective [8–9].

Beyond chronic disease management, research in advanced illness contexts demonstrates that empowerment is deeply experiential, involving identity, relational negotiation, and coping with losses, but it is rarely integrated into service design [10]. This further underscores the conceptual ambiguity of patient empowerment and the need to clarify how empowerment is experienced by patients themselves.

Taken together, these findings highlight uncertainty regarding which intervention components are most valued by patients and how they contribute to outcomes. Addressing this gap requires understanding how empowerment is experienced in everyday CHF management and how interventions can be optimally designed for specific health system contexts, such as Singapore.

This protocol describes the methods for a study to identify a patient-centred, empowerment-based intervention for adults with CHF that is both contextually appropriate and cost-effective in Singapore.

## METHODS AND ANALYSIS

We will adopt a staged, sequential design (Figure 1). First, the systematic review will synthesise international evidence on the effectiveness of empowerment-based interventions for adults with CHF. Next, as part of a mixed-methods study, we will conduct qualitative interviews with CHF patients and caregivers to explore how empowerment is experienced in local CHF care. We will then integrate insights from the systematic review and qualitative interviews to generate attributes and levels for a discrete choice experiment (DCE); the DCE will elicit patients’ preferences for features of an empowerment-based intervention and help identify a locally acceptable and implementable intervention package. In addition, the DCE will provide estimates of likely intervention uptake and adherence, which will inform the subsequent cost-effectiveness model. Finally, the intervention package selected based on the combined evidence from the systematic review, qualitative interviews and DCE will be evaluated in a cost-effectiveness analysis, comparing it with usual care from the Singapore healthcare system perspective to estimate costs, health outcomes, and overall value for money; where findings from the systematic review, qualitative interviews and DCE diverge, discrepancies will be considered in light of intervention effectiveness, cultural appropriateness, implementation feasibility and patient preferences when selecting the final intervention package.

**Figure 1.**
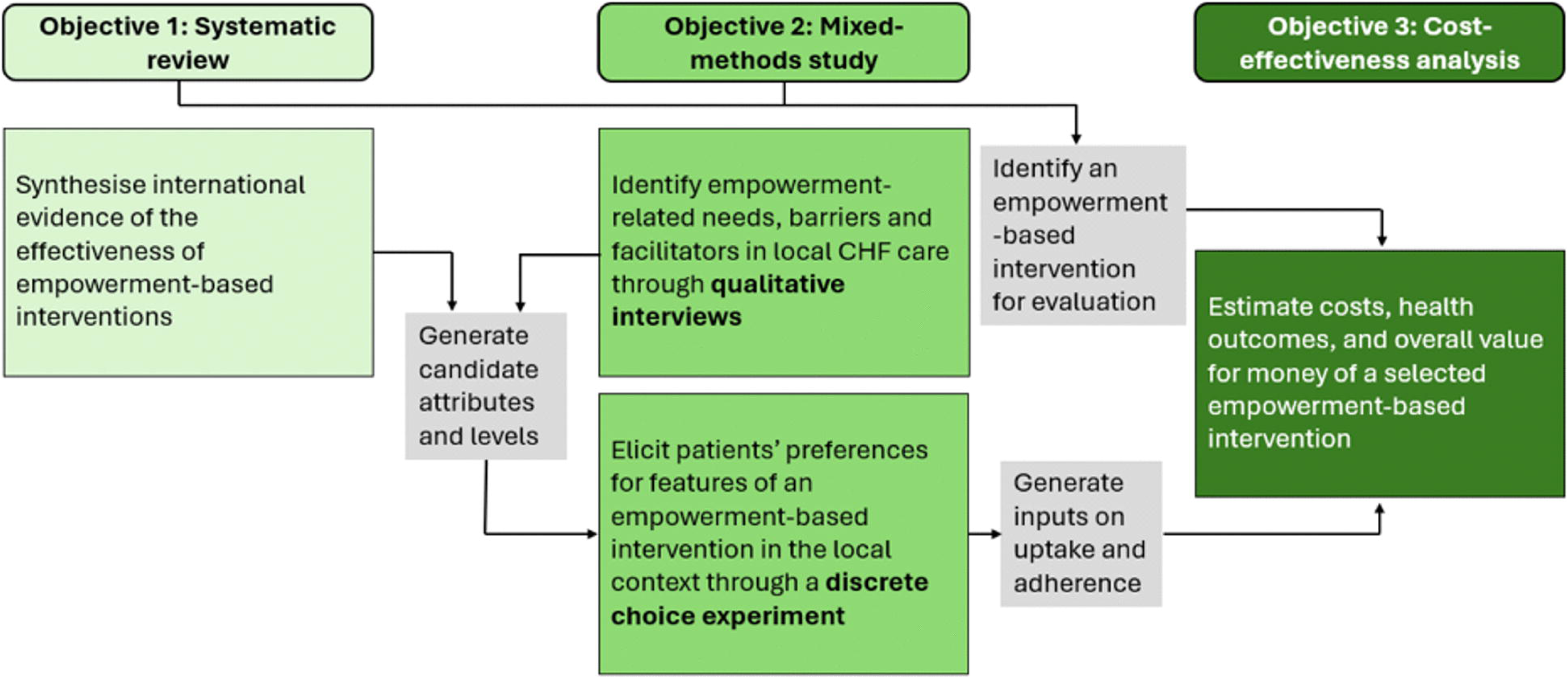
Depiction of the study’s staged sequential design.

### Objective 1 – Systematic review

#### Study registration

This systematic review was registered with PROSPERO (registration number CRD420251249957; http://www.crd.york.ac.uk/PROSPERO).

#### Research question

Despite increasing interest in empowerment-based approaches for CHF, no recent synthesis has comprehensively evaluated their effects. The review aims to address the following research question: Among adults with CHF, do interventions that use an empowerment approach, compared with (i) usual care, (ii) non-empowerment interventions, or (iii) other empowerment-based interventions (i.e., head-to-head comparisons), improve patient-reported outcomes (e.g., quality of life, symptoms), functional outcomes (e.g., exercise capacity), clinical outcomes (e.g., hospitalisations, mortality), and health service-related outcomes (e.g., healthcare costs) in the short and long term? This research question aligns with the first objective of this study, which is to determine the effectiveness of interventions that use an empowerment approach for patients with CHF.

#### Eligibility criteria

The review will be conducted and reported in accordance with the Preferred Reporting Items for Systematic Reviews and Meta-Analyses (PRISMA) guidelines [11]. Supplementary Material 1 contains the PRISMA-P checklist. Table 1 outlines the eligibility criteria of studies included in the review. We draw on the World Health Organisation’s (WHO) patient empowerment framework [12] to inform the definition of empowerment interventions in this review, which conceptualises empowerment as comprising four components: patient participation, patient knowledge, patient skills, and a facilitating environment.

**Table 1.**
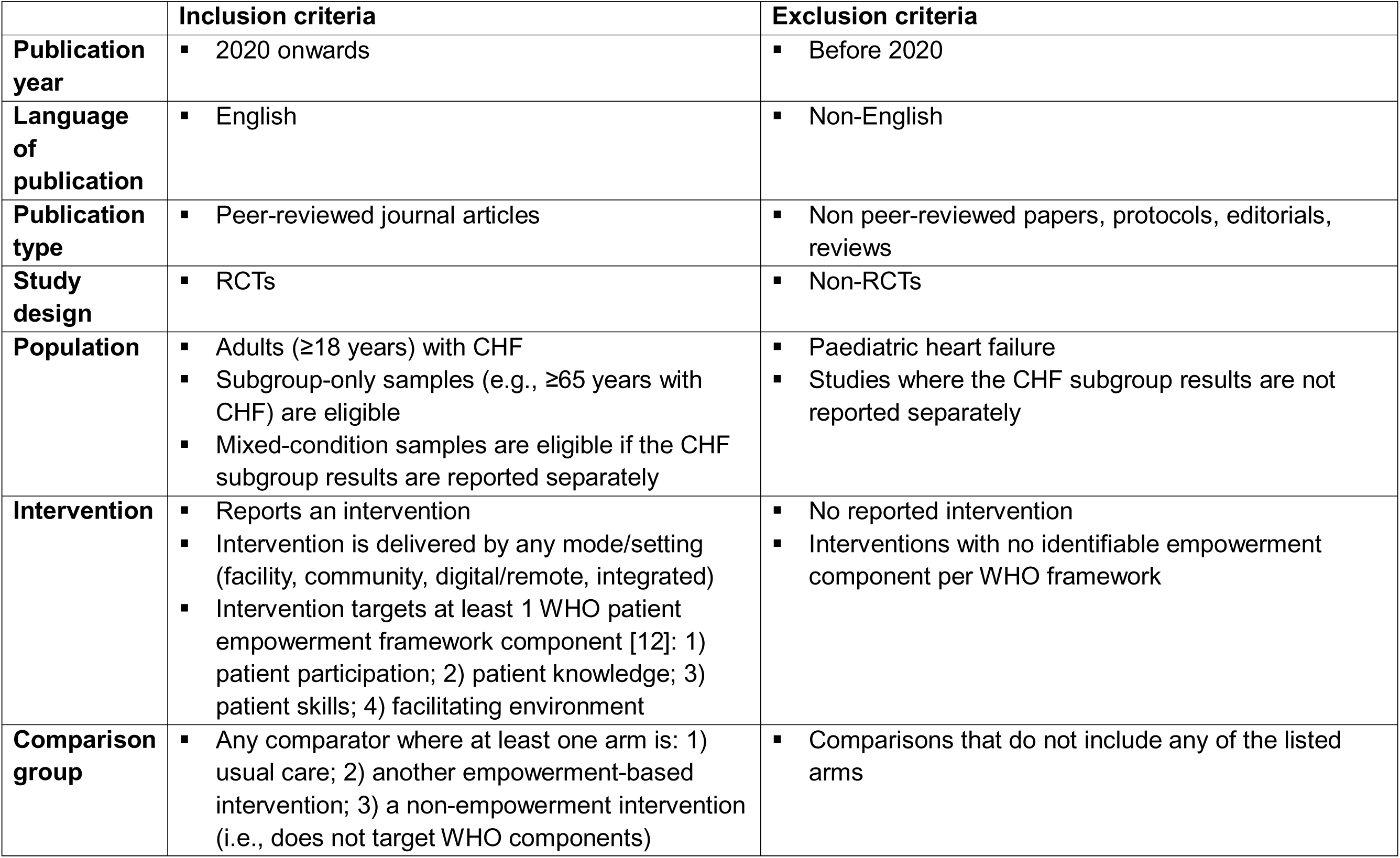
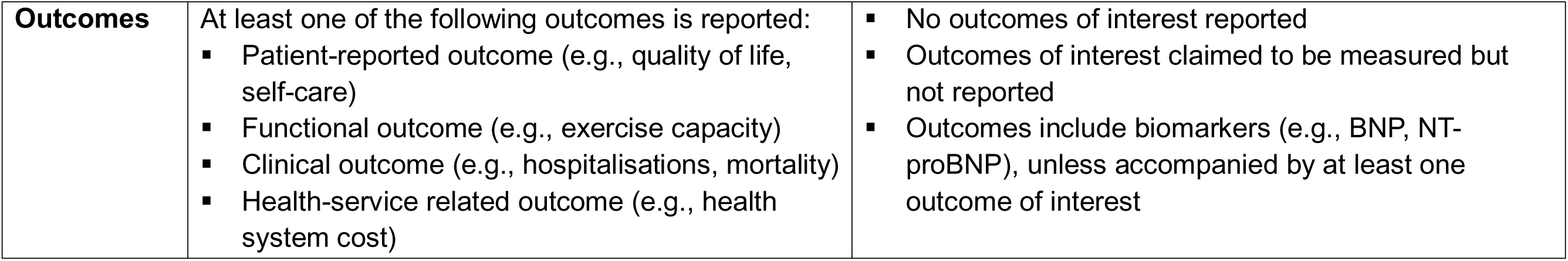
Study eligibility criteria.

|  | **Inclusion criteria** | **Exclusion criteria** |
| --- | --- | --- |
| **Publication year** | 2020 onwards | Before 2020 |
| **Language of publication** | English | Non-English |
| **Publication type** | Peer-reviewed journal articles | Non peer-reviewed papers, protocols, editorials, reviews |
| **Study design** | RCTs | Non-RCTs |
| **Population** | Adults ($\geq 18$ years) with CHF<br>Subgroup-only samples (e.g., $\geq 65$ years with CHF) are eligible<br>Mixed-condition samples are eligible if the CHF subgroup results are reported separately | Paediatric heart failure<br>Studies where the CHF subgroup results are not reported separately |
| **Intervention** | Reports an intervention<br>Intervention is delivered by any mode/setting (facility, community, digital/remote, integrated)<br>Intervention targets at least 1 WHO patient empowerment framework component [12]: 1) patient participation; 2) patient knowledge; 3) patient skills; 4) facilitating environment | No reported intervention<br>Interventions with no identifiable empowerment component per WHO framework |
| **Comparison group** | Any comparator where at least one arm is: 1) usual care; 2) another empowerment-based intervention; 3) a non-empowerment intervention (i.e., does not target WHO components) | Comparisons that do not include any of the listed arms |
| **Outcomes** | At least one of the following outcomes is reported:<br>▪ Patient-reported outcome (e.g., quality of life, self-care)<br>▪ Functional outcome (e.g., exercise capacity)<br>▪ Clinical outcome (e.g., hospitalisations, mortality)<br>▪ Health-service related outcome (e.g., health system cost) | ▪ No outcomes of interest reported<br>▪ Outcomes of interest claimed to be measured but not reported<br>▪ Outcomes include biomarkers (e.g., BNP, NT-proBNP), unless accompanied by at least one outcome of interest |

#### Search strategy

We will search four databases, namely, PubMed, Embase, Web of Science, and CINAHL, using database-tailored strategies that combine controlled vocabulary (MeSH/Emtree/CINAHL Subject Headings) and free-text keywords for the population and empowerment-related constructs. Across databases, we will apply limits for language (English), publication year (2020 onwards), human studies, and peer-reviewed empirical designs, reflecting contemporary CHF care and feasibility constraints. Full search strings and all limits are documented in Supplementary Material 2. Records will be deduplicated across databases. We will identify additional studies by screening the reference lists of included studies and published systematic reviews with a similar scope.

#### Study selection

We will screen deduplicated records manually, without the use of artificial intelligence, but with the assistance of the software, Rayyan, for organisation. Prior to screening, two reviewers will independently screen a random sample of 100 titles and abstracts to calibrate the eligibility criteria; conflicting decisions will be resolved, and the eligibility criteria will be refined accordingly. All remaining records’ titles and abstracts will then be independently screened by two reviewers, with a total of four reviewers. Conflicts will be reconciled within each pair, with involvement by a third reviewer if needed. For full-text screening, two reviewers will initially screen a random subset of records independently to calibrate the eligibility criteria. We will then split the remaining records between the two reviewers for independent screening. Any uncertainties or borderline cases will be discussed and resolved by consensus, with a third reviewer consulted if required.

#### Data extraction

We will design and pilot a standardised data extraction form on 5-10 studies, then have one reviewer independently apply it to all included studies. As data extraction progresses, we may further refine the data extraction form as required. Study characteristics (e.g., country, number and breakdown of participants), reported intervention features (e.g., delivery mode, dose and intensity, resources), and outcome data for patient-reported, functional, clinical, and health service-related outcomes will be extracted; no formal prioritisation of outcomes is planned. A second reviewer will independently extract data from a 10% random subset of records to verify completeness and accuracy of extraction.

#### Risk of bias assessment

We will use the Revised Cochrane Risk-of-Bias tool for randomised trials (Cochrane RoB-2) [13] to assess the risk of bias in all included studies. A single overall risk-of-bias judgement will be assigned to each study based on outcomes relevant to the review question. We will perform sensitivity analyses, where appropriate, excluding studies assessed as having a high risk of bias to assess the robustness of the results.

#### Analysis

We will perform meta-analyses for each outcome, where at least two studies report comparable data. We will calculate pooled effect estimates for dichotomous outcomes using risk ratios or hazard ratios, as appropriate, and for continuous outcomes using mean differences or standardised mean differences depending on whether consistent measurement instruments are used across studies. A random-effects model will be applied, given the anticipated clinical and methodological heterogeneity across empowerment-based interventions. We will assess statistical heterogeneity using the I² statistic and assess the potential for publication bias where sufficient studies are available for meta-analysis.

Where sufficient data are available, subgroup analyses will explore potential sources of heterogeneity by intervention characteristics (e.g., empowerment component, mode of delivery, dose and intensity). Where statistical pooling is not appropriate, we will synthesise findings narratively and structure them around predefined outcome categories (i.e., patient-reported outcomes, functional outcomes, clinical outcomes, and health-service related outcomes).

We will assess the certainty of the body of evidence for each outcome using the Grading of Recommendations Assessment, Development and Evaluation (GRADE) approach [14].

### Objective 2 – Mixed methods study: Qualitative study

#### Overview

The qualitative component aims to identify types of empowerment-based interventions that are likely to be effective and acceptable in the local context, and to understand how patients and caregivers perceive and experience empowerment in CHF care. We have obtained ethics approval through the SingHealth Centralised Institutional Review Board (ECOS Ref: 2025-1152).

#### Data collection procedures

We will use one-to-one, semi-structured interviews to explore patients’ and caregivers’ experiences and perspectives on empowerment in CHF management in Singapore. Semi-structured interviews are chosen as they allow in-depth exploration of patients’ and caregivers’ perceptions, needs, and barriers around empowerment-based CHF care, while maintaining enough structure to ensure coverage of key domains, and flexibility to probe context-specific issues as they arise [15–17]. Interviews will be conducted by trained research staff and will last approximately 45-60 minutes. Interviews may be conducted in English, Mandarin, Malay, or Tamil, either in-person or virtually, according to participant preference. With consent, interviews will be audio recorded to support transcription. Interview questions for patients and caregivers will cover topics relating to the four WHO empowerment components [12], and will include key information needs, self-management routines, appointment adherence and care navigation, communication with healthcare providers and shared decision-making, social support and caregiving roles, and cultural and religious considerations, among others (see Supplementary Materials 3 and 4 for interview questions). Interview questions will be piloted with initial participants and iteratively refined to enhance clarity and relevance.

#### Participants

Two participant groups will be enrolled: adult patients living with CHF and caregivers of patients with CHF. Eligible patients are Singapore citizens or permanent residents aged 21 years or older with a diagnosis of chronic heart failure, New York Heart Association (NYHA) class II or above [18], who can communicate in English, Mandarin, Malay, or Tamil and provide informed consent. Caregivers will comprise adults aged 21 years or older who provide care to a person with chronic heart failure (NYHA class II or above) and who can communicate in English, Mandarin, Malay, or Tamil and provide informed consent. Patients or caregivers with severe cognitive impairment or severe hearing or speech impairment that would preclude meaningful participation will be excluded.

We aim to recruit 30-60 patients and 30-45 caregivers, and will use purposive sampling to ensure variation by age, sex, language, and socioeconomic status (SES). SES will be captured via a brief screening questionnaire to support a balanced sample (see Supplementary Material 5). We will continue recruitment until adequate data are collected, with data adequacy primarily guided by the concept of information power, taking into account the study aim, sample specificity, use of an established theoretical framework, and analytic strategy [19]. Saturation will be used as a supporting indicator of the required sample size [20], reflected by a stabilisation of coding within the WHO-aligned components and the absence of substantive new insights.

Patients will be recruited through three medical institutions in Singapore, namely National University Hospital, Tan Tock Seng Hospital, and National Heart Centre Singapore, with targets of approximately 20-30 patients per institution. Recruitment will occur through attending healthcare providers at the outpatient clinics of these institutions, coordinated by study team members based in the respective institutions. Attending healthcare providers will introduce the study to eligible patients during their outpatient visits and, if interested, share a recruitment flyer with study details. Once patients are recruited, with the patient’s consent, their caregiver will be invited to participate. We will screen all interested individuals against the participant eligibility criteria before an interview is scheduled.

#### Data analysis

Interviews will be transcribed, and non-English interviews will be transcribed and translated into English by bilingual research staff. Anonymised transcripts will be analysed using codebook thematic analysis [21–22]. In this study, we will adopt a hybrid deductive-inductive approach, with the initial coding framework informed deductively by the four WHO empowerment components [12]. These components will serve as overarching themes, within which specific codes will be developed inductively from participants’ responses. NVivo will be used to facilitate data management and analysis.

To ensure coding reliability, a first coder will establish a preliminary coding framework based on an initial subset of transcripts (approximately 10-20%). A second coder will independently code this subset using the preliminary coding framework to calibrate the application of codes, after which the coding scheme may be refined through consensus. The remaining transcripts will then be coded by the first coder, with the coding framework further refined iteratively as needed. We will conduct regular team debriefs to discuss coding decisions and difficulties.

The final coding scheme will holistically reflect key empowerment-related experiences, needs, barriers and facilitators relevant to patients with CHF in Singapore, including aspects of care that participants perceive as meaningful, acceptable, and feasible in the local context.

#### Ensuring rigour

Rigour in the qualitative study will be supported through several strategies, including the piloting and refinement of interview questions, checks of translated transcripts, ensuring reliability in coding, and the maintenance of an audit trail to document rationale for decisions or changes made over the course of the study [23]. We will also practice reflexivity to reflect on and disclose how researcher positionality and potential biases may have affected data collection and analysis [24].

#### Conceptual framework

Findings from the systematic review and qualitative study will be synthesised into a conceptual framework to guide attribute selection and level specification for the DCE. The framework will diagrammatically represent: (1) hypothesised relationships among attributes (e.g., complementarity, mutual exclusivity, dominance) that may require bundling, conditional levels, or design constraints – for instance, when one attribute deterministically implies the other, we will avoid double counting by including only one; (2) respondent characteristics and contextual factors (e.g., heart failure classification, sex, care setting) that are not part of the alternatives but may moderate preferences and be used to explore heterogeneity in analysis; and (3) potential interactions between socio-demographic variables and intervention attributes. The framework will also record expected non-compensatory decision rules to guide the experimental design (e.g., constraints, blocking, split samples).

### Objective 2 – Mixed methods study: Discrete choice experiment

A DCE will be conducted to quantify CHF patients’ preferences for key attributes of empowerment-based interventions [25–27]. Figure 2 shows how this DCE will be developed, from identifying and selecting attributes and levels, through experimental design, pre-testing and piloting, final survey administration, econometric model, and data analysis. Stage 1 includes the systematic review and semi-structured interviews mentioned in the previous sections. Detailed plans for conducting Stages 2 to 4 will be presented in this section.

**Figure 2.**
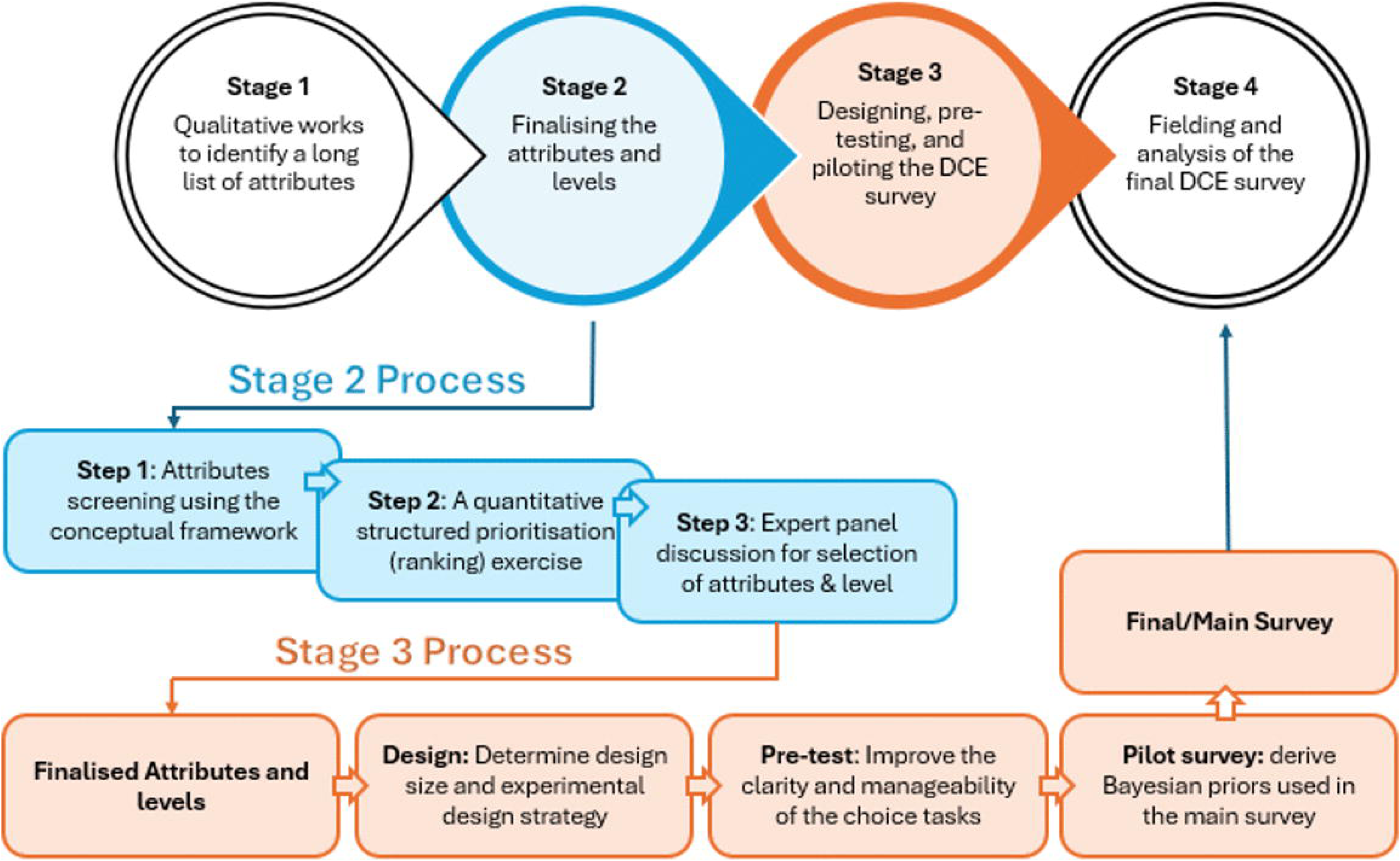
Discrete choice experiment development procedure.

#### Stage 2: Finalising the attributes and levels

The initial selection of attributes and levels will be informed by the findings of the systematic review and the qualitative study [28–29]. We will use an unlabelled DCE with two or three generic intervention alternatives, as this best aligns with our aim of eliciting preferences for intervention features. An opt-out option will be included consistently in each choice task to reflect realistic decision contexts and to anchor the interpretation of choices. The following three steps will be taken to finalise the attributes and the corresponding levels.

##### Step 1

The selected candidate attributes will first be screened using criteria informed by the conceptual framework. We will retain attributes that are decision-relevant and salient to patients, and that can realistically vary and be acted upon in service delivery or policy. We will also assess whether each attribute is genuinely tradeable and exclude absolute “must-haves” attributes for most respondents. In addition, we will examine the relationships between attributes to identify overlaps or deterministically linked constructs to merge or exclude redundant attributes. “Features of the intervention” and “respondent/context factors” will be distinguished, with the latter excluded or held constant as fixed scenario information in the choice tasks.

##### Step 2

Using the reduced attribute list, we will conduct a quantitative, structured prioritisation exercise to assess the relative importance of the essential attributes from the patient perspective. Eligible heart failure patients will complete a brief survey where they rank attributes from “most” to “least” important. Rankings will be converted to scores, with higher rankings corresponding to lower scores, and the mean score will also be calculated for each attribute. These scores will be used to inform expert panel discussions to determine a much smaller list of attributes and associated levels.

##### Step 3

Attributes identified as of greater importance will form the basis of discussion and finalisation by the expert panel comprising health economists, scientific researchers, clinicians involved in heart failure research, and patient representatives. The panel will consider the following factors when deciding the finalised attributes for the DCE survey: 1) patient inputs from Step 2 about the relative importance; 2) relevance of attributes to the goal of the DCE; and 3) significance and applicability of the attributes.

Once the attribute set is agreed upon, the panel will discuss the attribute levels. The number of levels per attribute will be kept manageable and balanced across attributes where possible. If a cost attribute is included, its perspective will be defined explicitly to support meaningful marginal rates of substitution, e.g., willingness-to-pay. Meanwhile, the scenario to be presented to the survey respondents will be developed. After the discussion, meeting notes and the proposed attribute list with levels will be reviewed, refined, and finalised.

#### Stage 3: Designing, pre-testing, and piloting the DCE Survey

##### Design

With the final attribute–level set, we will construct a D-efficient fractional factorial design using Ngene [30]. The theoretical minimum number of choice tasks required to estimate all parameters is defined by the number of attribute levels/(number of alternatives -1) [31]. In practice, substantially larger designs – often around two to three times this minimum – will be applied to improve statistical efficiency and design quality. To manage respondents’ cognitive burden, the choice tasks will be equally partitioned into several blocks, with each block containing no more than 10 tasks.

##### Pre-test

Pre-testing will include expert review (clinical and methodological) and cognitive interviews with a small, purposive sample of patients to probe how they interpret the attributes/levels, the decision context, and the instructions, and to identify misunderstandings, ambiguous terms, and excessive cognitive burden.

Revisions will be made iteratively until the choice tasks, attribute wording, and level descriptions are comprehensible, salient, and feasible for respondents.

##### Pilot survey

We will then conduct a pilot to evaluate survey completion and obtain informative priors for the final experimental design. We will use the multinomial logit model to analyse the pilot survey data and to obtain pilot parameter estimates, which will be used to update Bayesian efficient priors [32–33] in Ngene and, where necessary, to re-optimise the design. In addition to the choice responses, we will collect structured feedback on clarity, burden, realism, and interface/usability to guide final adjustments before launching the main survey.

In addition to responses to the choice tasks, we will collect socio-demographic information, clinical heart failure profile, brief survey evaluation items, and indicators of the decision process, such as attribute non-attendance. These data will help explain preference heterogeneity, support subgroup and interaction analyses, diagnose potential scale differences, and lexicographic behaviour.

#### Stage 4: Fielding and analysis of the final DCE survey

##### Sample size determination

We will use the S-error output [34–35] from Ngene to determine the minimum sample size required to achieve acceptable statistical precision and increase this minimum to account for practical considerations so that the achieved sample size supports the desired precision in the final estimates.

##### Validity check

Apart from the finalised choice tasks, we will incorporate the following checks into the final survey to assess data quality: 1) a repeat choice task and a dominant choice task to check whether respondents are attending to the attribute information; 2) identify fast responders who complete the survey in an extremely short time; and 3) ask respondents to report their age early in the survey and their year of birth near the end, where discrepancies beyond an acceptable range will be flagged as potential inattention. Apart from guiding pre-specified data cleaning rules, these checks will also be embedded in sensitivity analyses to evaluate the robustness of the main findings.

##### Data analysis

We will pool data from the pilot and the main study and account for possible scale differences. We will estimate a Multinomial Logit (MNL) model for a primary analysis and to examine observed heterogeneity via attribute–covariate interactions with socio-demographic and clinical variables. We will then employ both the mixed Multinomial Logit (MMNL) and latent class models with covariates to describe unobserved preference heterogeneity. The model fit will be assessed subsequently. If a cost attribute is included, we will report willingness-to-pay (WTP) measures as marginal rates of substitution, and for MMNL/latent class models, we will present distributions or class-specific WTP to reflect heterogeneity. The relative importance of attributes will also be computed. Finally, we will conduct scenario analyses to predict the uptake of policy-relevant intervention packages relative to the opt-out option.

### Objective 3 – Cost-effectiveness analysis

#### Overview

Following the DCE, we will conduct a cost-effectiveness analysis to determine the cost-effectiveness of a patient empowerment intervention compared with usual care for CHF. The empowerment intervention selected for evaluation will be informed by findings from the preceding studies (systematic review, qualitative interviews, and DCE), as well as expert opinion.

This analysis will use a decision-analytic model combining a decision tree and a Markov state-transition model. A decision tree will model the initial allocation to intervention or usual care, followed by a Markov model to simulate long-term disease progression. We will develop the model using TreeAge Pro 2025. The target population comprises CHF patients of NYHA class II and above, with a median age of 60 years.

This study will be conducted based on a predefined economic evaluation plan, and findings will be reported in accordance with the Consolidated Health Economic Evaluation Reporting Standards (CHEERS) guidelines [36].

#### Model structure

The model consists of two components: a decision tree followed by a Markov state-transition model (Figure 3). The decision tree reflects the initial choice between Intervention and Usual care. Within the intervention arm, patients are further stratified by uptake and adherence, with the associated probabilities informed by the DCE. At the end of each decision-tree pathway, a Markov model with the same health-state structure is simulated.

**Figure 3.**
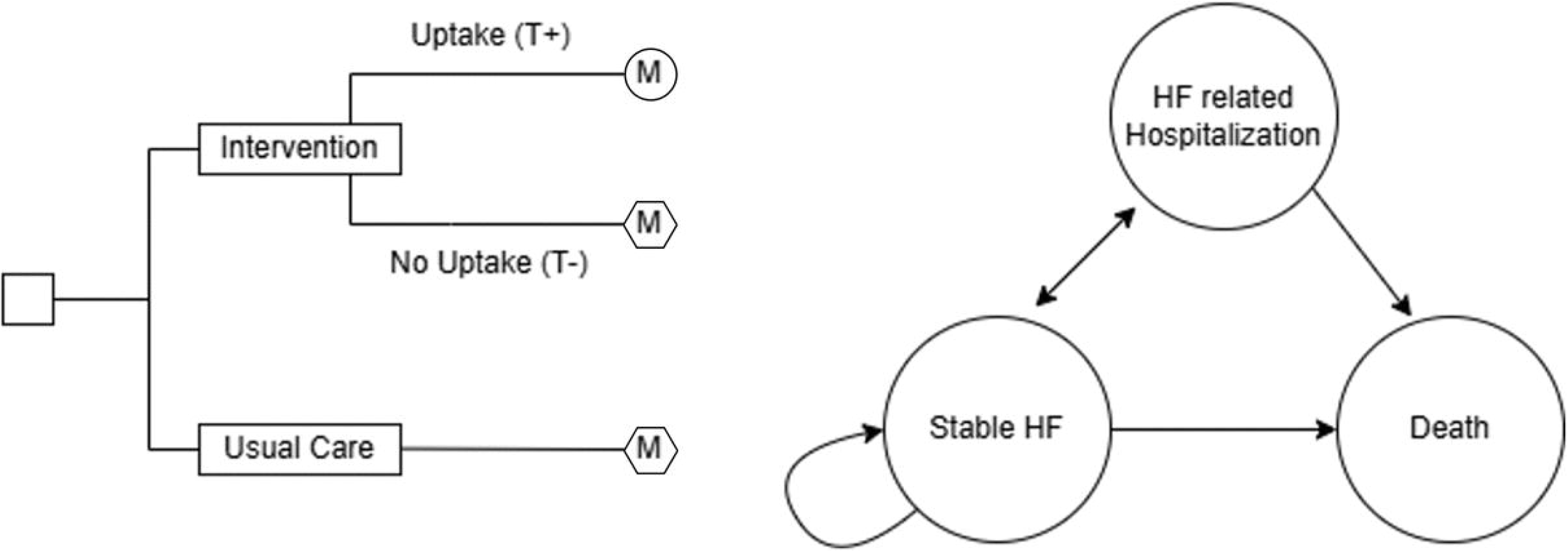
Decision Tree Pathway and the Three health states of the Markov state-transition model.

The Markov state-transition model consists of three mutually exclusive health states: Stable Heart Failure (clinically stable patients of NYHA class II and above), HF related hospitalisation (inpatient episode due to CHF), and Death (absorbing state).

All patients will enter the model in the Stable Heart Failure (HF) state. During each cycle, patients may remain stable, experience hospitalisation, or die. Patients who are hospitalised may transition back to the Stable Heart Failure state following discharge or die. Death is an absorbing state with no further transitions.

The Markov models for the Usual Care arm and for patients who do not uptake the intervention are identical (hexagon Markov symbol in Figure 3), reflecting the same baseline transition probabilities. In contrast, patients who receive and adhere to the intervention are modelled with intervention-specific transition probabilities (circle Markov symbol in Figure 3) to capture anticipated improvements in outcomes, such as lower risks of HF-related hospitalisation or mortality.

The analysis will adopt a Singapore health system perspective over a 10-year time horizon with annual cycles. Effectiveness will be measured in quality-adjusted life years (QALYs) and compared against total accumulated costs. Both QALYs and costs will be discounted at an annual rate of 5%.

#### Data sources

We will derive time-dependent transition probabilities for the usual-care arm from Senanayake et al. (2026) [37], a Singapore-based cost-effectiveness study of improving guideline-directed medical therapy in heart failure. Senanayake et al. [37] report time-dependent probabilities (parameterised using Weibull regression) for the following transitions: Stable HF to HF related hospitalisation, stable HF to death, and HF related hospitalisation to death. We will use the reported Weibull parameters (scale *λ* and *γ* shape) to generate cycle-specific transition probabilities for each annual Markov cycle. For the intervention arm, we will apply the intervention effect estimate (e.g., hazard ratio for hospitalisation and/or mortality), obtained from the selected clinical trial evidence or the meta-analysis in Objective 1, to the corresponding baseline hazards before converting them into cycle-specific transition probabilities.

Costs associated with each health state (Stable HF, and HF related hospitalisation) will be informed by the same Singapore-based study by Senanayake et al. (2026) [37], which reports state-specific costs from the Singapore healthcare system perspective. We will obtain the cost of the selected empowerment intervention from published literature where available; if not available, we will estimate it using local resource-use assumptions informed by clinical and operational experts and implemented through scenario-building.

As local health utility data are unavailable, regional and international estimates will be used to assign utility weights to each health state. The mean utility values are 0.71 (SD 0.016) for the Stable Heart Failure state [38] and 0.605 (SD 0.017) for the heart-failure-related hospitalisation state [39].

#### Model evaluation and interpretation

Upon model simulation, total accumulated costs and QALYs will be estimated for both the usual care and intervention arms, and incremental costs and QALYs will be calculated to derive the incremental cost-effectiveness ratio (ICER). The ICER will be compared against a predefined willingness-to-pay (WTP) threshold of $45,000 per QALY, reflecting thresholds previously applied in the Singapore context [40], to determine cost-effectiveness. We will conduct probabilistic sensitivity analyses to assess the impact of parameter uncertainty on model outcomes. Monte Carlo simulation with 1,000 iterations will be performed, applying beta distributions for utilities, Weibull distributions for transition probabilities and gamma distributions for costs. We will generate a cost-effectiveness acceptability curve (CEAC) to illustrate the probability that the intervention is cost-effective across a range of WTP values.

Scenario analyses will further explore structural and assumption uncertainty, including alternative assumptions regarding intervention effect, uptake, delivery models, and discount rates. Net monetary benefit (NMB) will be estimated for each scenario by valuing health gains at the specified WTP threshold and subtracting incremental costs. The scenario with the highest NMB will be considered the most economically favourable.

#### Patient and public involvement

Patients were not involved in developing the research question or in the design of the systematic review or cost-effectiveness analysis. Patient representatives with lived experience of CHF will sit on the expert panel that finalises the DCE attributes and levels (Objective 2, Stage 2).

#### Status and timeline of study

The study is ongoing. For the systematic review component, data extraction is expected to be completed in December 2026, with findings anticipated in February 2027. For the remaining study components, participant recruitment began in March 2026 and is ongoing; recruitment and data collection are expected to be completed in January 2027. The overall study is expected to be completed in March 2027, with final study findings anticipated at that time.

## ETHICS AND DISSEMINATION

### Ethical considerations

The systematic review and cost-effectiveness analysis use published and aggregate data and do not require ethics approval. Ethics approval for the qualitative study has been obtained from the SingHealth Centralised Institutional Review Board (ECOS Ref: 2025-1152). Ethics approval for the DCE study will be submitted prior to participant recruitment. Qualitative study participants interviewed in person will provide written informed consent, while those interviewed online will provide verbal consent, which will be audio recorded for documentation. Participation is voluntary, and participants may withdraw at any time without penalty.

### Safety and confidentiality

The study involves no clinical procedures and minimal risks to participants. Participants may experience emotional discomfort when discussing their illness or caregiving during the qualitative interviews; however, they will be informed of their ability to skip any question or stop the interview at any time. Data will be de-identified, with the code key stored separately. All data will be stored on encrypted, institution-approved storage accessible only to the study team.

### Dissemination

Findings from each study component will be submitted for publication in peer-reviewed journals and presented at national and international conferences.

### Data deposition and curation

Systematic review search strategies are provided in the supplementary material, and the extracted data will be published with the review. Qualitative data will not be shared, to protect participant confidentiality. The DCE survey instrument and experimental design will be published as supplementary material, and cost-effectiveness model inputs will be reported in accordance with CHEERS 2022. De-identified DCE data and the cost-effectiveness model will be available from the corresponding author on reasonable request.

## Data Availability

All relevant data from this study will be made available upon study completion.

## AUTHOR CONTRIBUTIONS

ALSY, RCCW, SYL, NG, SSu, SY, DJH, LLL, KKY, KLDS, YZ, SK and SSe contributed to conceiving the study. SK, SSe, BV, YW and DKR contributed to the design of the study methods. BV, YW and DKR drafted the manuscript. All authors critically revised the manuscript for important intellectual content, approved the final version, and agree to be accountable for all aspects of the work. SK and SSe contributed equally as co-senior authors. SK is the guarantor.

