## Supplementary Material 2 for "Empowering Singaporeans to better manage chronic heart failure: Protocol for a mixed-methods study"

**Supplementary Material 2. Search strategy documentation.**

| **Medline (PubMed)** | **Controlled vocabulary** **terms**  MeSH terms | Heart Failure | | Patient participation   - [MeSH] Patient participation - [Mesh Major Topic] Consumer participation   Patient knowledge   - [MeSH] Patient education as topic - [MeSH] Health education - [MeSH] Counseling   Patient skills   - [MeSH] Self care - [MeSH] Self-management - [MeSH] Self efficacy - [MeSH] Health literacy   Facilitating environment   - [MeSH] Patient-centered care - [MeSH] Decision making, shared | |
| --- | --- | --- | --- | --- | --- |
|  | **Free text keywords** | Heart failure OR Cardiac failure | | Patient participation   - Empower* - Patient participat* - Patient engag* - Patient activation - Motivational interview*   Patient knowledge   - Patient educat* - Consumer educat*   Patient skills   - Self-manag* - Self manag* - Self-care - Self care - Self-efficacy - Self efficacy* - Health literacy*   Facilitating environment   - Patient-provider communication - Physician-patient communication - Shared decision making - Shared decision* - Positive devian* | |
|  | **Search string** | ("Heart Failure"[mh] OR "heart failure"[tiab] OR "cardiac failure"[tiab]) AND ("Patient Participation"[mh] OR "Consumer Participation"[majr] OR "Patient Education as Topic"[mh] OR "Health Education"[mh] OR "Counseling"[mh] OR "Self Care"[mh] OR "Self-Management"[mh] OR "Self Efficacy"[mh] OR "Health Literacy"[mh] OR "Patient-Centered Care"[mh] OR "Decision making, shared"[mh] OR empower*[tiab] OR patient participat*[tiab] OR patient engag*[tiab] OR "patient activation"[tiab] OR motivational interview*[tiab] OR patient educat*[tiab] OR consumer educat*[tiab] OR self-manag*[tiab] OR self manag*[tiab] OR "self-care"[tiab] OR "self care"[tiab] OR "self-efficacy"[tiab] OR self efficacy*[tiab] OR health literacy*[tiab] OR "patient-provider communication"[tiab] OR "physician-patient communication"[tiab] OR shared decision*[tiab] OR "shared decision making"[tiab] OR positive devian*[tiab]) | | | |
|  | **Limits/filters** | - 2020 onwards - Published in English - Humans - Exclude preprints - Study type: Clinical Trial, Comparative Study, Controlled Clinical Trial, Evaluation Study, Observational Study, Pragmatic Clinical Trial, Randomized Controlled Trial, Validation Study | | | |
| **Embase** | **Controlled vocabulary** **terms**  Emtree terms | Heart Failure | | | Patient participation   - Patient empowerment/ exp - Patient participation/ exp   Patient knowledge   - Patient education/ exp   Patient skills   - Self care/ exp   Facilitating environment   - Shared decision making/ exp |
|  | **Free text keywords** | Heart failure OR Cardiac failure | | | Patient participation   - Empower* - Patient participat* - Patient engag* - Patient activation - Motivational interview*   Patient knowledge   - Patient education - Patient educat* - Health education - Counseling   Patient skills   - Self-care - Self care - Self-manag* - Self management - Self-efficacy - Self efficacy* - Health literacy   Facilitating environment   - Shared decision making - Shared decision* - Patient-provider communication - Physician-patient communication - Communication skills - Positive devian* - Positive deviance intervention |
|  | **Search string** | ('heart failure'/exp OR ("heart failure" OR "cardiac failure"):ti) AND ('patient empowerment'/exp OR 'patient participation'/exp OR 'patient education'/exp OR 'self care'/exp OR 'shared decision making'/exp OR (empower* OR patient participat* OR patient engag* OR "patient activation" OR motivational interview* OR patient education OR patient educat* OR "health education" OR counseling OR self-manag* OR "self management" OR "self-care" OR "self care" OR "self-efficacy" OR self efficacy* OR "health literacy" OR "patient-provider communication" OR "physician-patient communication" OR "communication skills" OR shared decision* OR "shared decision making" OR positive devian* OR "positive deviance intervention"):ti,ab) | | | |
|  | **Limits/filters** | - 2020 onwards - Published in English - Publication types: Article, Article in Press | | | |
| **Web of Science** | **Free text keywords** | Heart failure OR Cardiac failure | Patient participation   - Empower* - Patient participat* - Patient empowerment - Patient activation - Motivational interview*   Patient knowledge   - Patient education - Health education   Patient skills   - Self-efficacy - Self efficacy* - Self-manag* - Self care - Self-care - Health literacy*   Facilitating environment   - Communication skills - Patient-provider communication - Physician-patient communication - Positive devian* - Positive deviance intervention | | |
|  | **Search string** | TI=("heart failure" OR "cardiac failure") AND TS=(empower* OR patient participat* OR "patient empowerment" OR "patient activation" OR motivational interview* OR "patient education" OR "health education" OR self-manag* OR "self care" OR "self-care" OR "self-efficacy" OR self efficacy* OR health literacy* OR "patient-provider communication" OR "physician-patient communication" OR "communication skills" OR positive devian* OR "positive deviance intervention") | | | |
|  | **Limits/filters** | - 2020 onwards - Published in English - Document Types: Article or Early Access | | | |
| **CINAHL** | **Controlled vocabulary** **terms**  CINAHL Subject Headings | Heart Failure | | | Patient participation   - [MH] Empowerment - [MH] Patient participation   Patient knowledge   - [MH] Patient education   Patient skills   - [MH] Self-Efficacy - [MH] Decision support techniques   Facilitating environment   - [MH] Professional-patient relations - [MH] Physician-patient relations - [MH] Nurse-patient relations |
|  | **Free text keywords** | Heart failure OR Cardiac failure | | | Patient participation   - Empower* - Patient participat* - Patient engag* - Patient activation - Motivational interview*   Patient knowledge   - Patient educat* - Counseling   Patient skills   - Self-manag* - Self care - Self-care - Self-efficac* - Health literacy - Decision aid*   Facilitating environment   - Patient-provider communication - Shared decision making - Positive devian* - Positive deviance intervention |
|  | **Search string** | (MH ("Heart Failure") OR XB ("Heart failure" OR "Cardiac failure")) AND (MH (Empowerment OR "Patient participation" OR "Patient education" OR "Self-efficacy" OR "Decision support techniques" OR "Professional-patient relations" OR "Physician-patient relations" OR "Nurse-patient relations") OR XB (empower* OR patient participat* OR patient engag* OR "patient activation" OR motivational interview* OR patient educat* OR counselling OR self-manag* OR "self care" OR "self-care" OR self-efficac* OR "health literacy" OR decision aid* OR "shared decision making" OR "patient-provider communication" OR positive devian* OR "positive deviance intervention")) | | | |
|  | **Limits/filters** | - 01/01/2020 onwards - Published in English - Peer reviewed - Human - Research Article | | | |
