## Supplementary Material 3 for "Empowering Singaporeans to better manage chronic heart failure: Protocol for a mixed-methods study"

**Supplementary Material 3. Patient interview questions.**

**Background**

1. Could you tell me a little about yourself?

Prompt: For example, your age, what you do day-to-day, or anything you’d like to share.

2. How long have you been living with your heart failure condition?
3. Can you tell me about your heart condition and the medication or treatment you are currently taking?
4. Are there any other health issues that you are currently managing alongside your heart condition?
5. Could you briefly describe your current living situation?

Prompt: Who do you live with? Do you live alone?

6. When you hear the word empowerment, or when you think about feeling in control of your health, what does that mean to you in relation to living with heart failure?

**Patient knowledge**

7. What kinds of knowledge or information do you think are most important for you when it comes to living with heart failure?

Prompt: Information about your medicines, how the condition may progress, or recognising worsening symptoms.

8. What information have you received about heart failure since your diagnosis?

Prompt: How clear was the information you got from your doctors or nurses? Have you been told what signs and symptoms to watch out for?

9. What sources of information have been most useful for you (e.g, healthcare team, internet, family, support groups)?
10. Have you attended any online courses or used online resources to learn about your health?

Prompt: How did you find that experience?

11. Do you feel you have enough information to make decisions about your treatment?
12. What kind of information would make you feel more in control of your health?

**Patient skills**

13. Can you walk me through a typical day of how you manage your heart condition?
14. How do you usually keep track of your health and symptoms?

Prompt:

- How regularly do you track any of your health aspects like weight, blood pressure or pulse?
- How do you track these measurements?
- Do you use any health devices (like blood pressure monitors, smart watches) to keep track of your health? Can you tell me more about your experience using them?

15. What do you do if you notice your symptoms getting worse?

Prompt: Do you feel confident that you can manage your symptoms if they get worse?

16. Do you follow advice on diet, sleep, exercise, medications, and lifestyle?

Prompt:

- How easy or difficult is it for you to follow these advice?
- If easy: How has it become easy; if difficult: Why has it become difficult?

17. What would make you feel more empowered to take care of yourself day-to-day?
18. How much control do you feel you have over keeping your heart failure stable?
19. Thinking about follow-up visits or tests, have you ever missed (or postponed) an appointment? If yes, can you share what happened?

Prompt:

- What were the main reasons? (e.g., symptoms, work/caregiving, transport, cost, scheduling, forgetfulness, clinic wait times, language, digital access).
- What would make it easier for you to attend or reschedule? (e.g., reminders, flexible hours, telehealth, transport support, shorter waits, clearer instructions).
- Who helps you keep appointments (family, caregivers), and what supports would help more?

20. When you miss or delay an appointment, how do you manage your heart failure in the meantime? Who do you contact?

**Facilitating environment**

21. How involved do you feel in decisions about your heart failure care and treatment?

Prompt: Is it mostly your doctor, family, or yourself?

22. What makes it easier or harder for you to take part in these decisions?

Prompt:

- Do you feel comfortable asking your healthcare providers questions or raising concerns?
- Have you ever felt your opinion wasn’t considered in decisions about your care?
- What kind of information or support would help you feel more confident in making decisions?

23. What does being part of decisions mean to you in feeling empowered?
24. What changes would make you feel more empowered in the way decisions about your treatment are made?
25. In what ways does communication with your healthcare team help you feel empowered?
26. Have you communicated with doctors, nurses, or other healthcare workers using phone calls, video calls, or messages?

Prompt:

- If yes, what was that like?
- If no, how do you usually communicate with them?

27. Who do you turn to for support in living with heart failure (family, friends, peers, healthcare team)? How do these people support you in managing your condition?
28. Do you talk to others (friends, people with the same condition, support groups) about your heart condition or how you feel? If yes, how does that make you feel?
29. Do you attend any counselling sessions? If yes, how do you find them?
30. Do you attend any peer support groups? If yes, how do you find those sessions?
31. How does having (or not having) support affect your confidence in managing heart failure?
32. Are there any cultural or religious factors that you consider when managing your heart condition? If yes, can you tell me more about that?

**Closing**

33. Do you feel confident managing your health condition?

Prompt: Why or why not? What would help you feel more confident?

34. What advice would you give to someone who is newly diagnosed with the same heart condition as you?
35. Apart from what we have discussed, would you like to share anything else?
