## Supplementary Material 4 for "Empowering Singaporeans to better manage chronic heart failure: Protocol for a mixed-methods study"

**Supplementary Material 4. Caregiver interview questions.**

**Background**

1. Can you tell me a little about yourself and your relationship to the person/loved one you care for?

Prompt: Your age, what you do, and your caregiving role.

2. How long have you been caring for your loved one with heart failure and how long does he/she have heart failure?
3. Can you tell me about the patient’s heart condition and the treatments or medications they are currently on?
4. Are there other health conditions your loved one has that you help manage?
5. Can you briefly describe your loved one’s living situation?

Prompt: Do you live together? Who else is in the household?

6. When you think about your loved one feeling “empowered” or “in control” of their health, what does that mean to you in relation to their heart failure?

Prompt: What does “empowerment” look like for them, from your perspective?

**Patient knowledge**

7. What kinds of knowledge or information do you think are most important for your loved one in managing heart failure?

Prompt: Information about medicines, condition progression, recognising symptoms, etc.

8. What information has your loved one received from healthcare professionals or others since their diagnosis?

Prompt: Was the information clear? Did you help explain or reinforce information?

9. What sources of information have been most useful for you and your loved one (e.g., healthcare team, internet, family, support groups)?
10. Has your loved one used any online resources or classes to learn about heart failure?

Prompt: What did you notice about their experience?

11. Do you feel your loved one has enough information to make decisions about their treatment?

Prompt: Do you ever need to look up information together?

12. What kind of information or resources would help your loved one—and you as caregiver—feel more in control?

**Patient skills**

13. Can you describe a typical day of how your loved one manages his/her heart condition?

Prompt: What is your role in supporting or reminding them?

14. How does your loved one keep track of health and symptoms (e.g., weight, blood pressure, pulse)?

Prompt: Who does the tracking? How often?

15. What do you or your loved one do if their symptoms get worse?

Prompt: Does your loved one feel confident in managing his/her symptoms? Do you feel confident in helping the patient manage worsening symptoms?

16. Does your loved one follow advice on diet, sleep, exercise, medication, or lifestyle?

Prompt: How easy or difficult is it for your loved one to follow this advice?

17. What do you think will help them feel more empowered to take care of themselves day-to-day?
18. How much control do you think your loved one feel they have over keeping their heart condition stable?
19. Thinking about your loved one’s follow-up visits or tests, have they ever missed (or postponed) an appointment? If yes, can you share what happened?

Prompts:

- What were the main reasons from your perspective? (e.g., their symptoms, your work/caregiving duties, transport, cost, scheduling, forgetfulness, clinic wait times, language, digital access)
- What would make it easier for them to attend or reschedule? (e.g., reminders, flexible hours, telehealth, transport support, shorter waits, clearer instructions)
- Who helps them keep appointments (family members, caregivers/you), and what supports would help more?

20. When an appointment is missed or delayed, how is your loved one’s heart failure managed in the meantime? Who do you or they contact?

**Facilitating environment**

21. How involved is your loved one in the decisions about their care and treatment?

Prompt: Are the decisions usually made by the patient, family, or healthcare team?

22. What makes it easier or harder for them to take part in these decisions?

Prompt: Are there times when you need to advocate for them or step in?

23. What does being part of decisions mean for them in the sense of empowerment, from your point of view?
24. What changes could help them feel more empowered during treatment decisions?

Prompt: What changes could help you as a caregiver?

25. In what ways does communication with healthcare professionals help your loved one feel empowered?

Prompt: Are there ways it could be improved?

26. Have you seen your loved use any type of technology—like phone calls, video calls, apps, or devices—to talk with healthcare professionals or look after their health?

Prompt: What have you noticed about how they get along with these tools?

27. Who provides support to your loved one?

Prompt: Family, friends, peers, healthcare team, etc. How do you see your role?

28. Does your loved one talk to others about their condition or feelings (e.g., friends, other patients, support groups)?

Prompt: If yes, how does this seem to affect them?

29. Does he or she attend any counselling or support group sessions?

Prompt: If yes, how do you think they find these experiences?

30. How does having (or not having) support affect your loved one’s confidence in managing heart failure?
31. Are there any emotional, cultural, or religious factors you’ve noticed that affect how your loved one manages their heart condition or asks for support?

**Closing**

32. From what you’ve seen, is there anything that you think is missing or any extra support that would really help your loved one?
33. What kinds of support, programs, or advice do you think would make it easier for your loved one to manage their heart condition?
34. Is there anything else you’ve observed or would like to share that we haven’t talked about today?
