## Supplementary Material 5 for "Empowering Singaporeans to better manage chronic heart failure: Protocol for a mixed-methods study"

**Supplementary Material 5. SES screening questionnaire.**

| 1. Title | ( ) Mr. ( ) Ms. ( ) Miss. ( ) Mrs. ( ) Mdm. ( ) Dr. |
| --- | --- |
| 2. Your Name: |  |
| 3. Your Contact Number: |  |
| 4. Your Email Address: |  |
| 5. Please tick the option that applies: | ( ) I have been diagnosed with heart failure by a physician.  ( ) I am a caregiver of someone who has been diagnosed with heart failure by a physician.  ( ) Neither of the above. |
| 6. If you are a **heart failure patient**, please tick all that apply:   ( ) I am a Singapore citizen or permanent resident aged 21 and above.  ( ) I been diagnosed with heart failure by a physician, classified as New York Heart Association (NYHA) Class II or above.  ( ) I am able to communicate in either English, Mandarin, Malay, or Tamil.  ( ) I do not have any severe cognitive, hearing or speech impairment. | If you are a **caregiver of a heart failure patient**, please tick all that apply:  ( ) I am an adult aged 21 and above.  ( ) I am caring for someone who has been diagnosed with heart failure by a physician, classified as New York Heart Association (NYHA) Class II or above  ( ) I am able to communicate in either English, Mandarin, Malay, or Tamil.  ( ) I do not have any severe cognitive, hearing or speech impairment. |
| 7. Your Age | ( ) 21-24  ( ) 25-34  ( ) 35-44  ( ) 45-54  ( ) 55-64  ( ) 65 and above |
| 8. Your Education Level | ( ) No formal education ( ) Primary school ( ) Secondary school ( ) Institute of Technical Education (ITE)/Vocational training ( ) Diploma/Polytechnic ( ) University degree ( ) Postgraduate degree ( ) Prefer not to say |
| 9. Your Housing Type | ( ) HDB 1–2 room flat ( ) HDB 3 room flat ( ) HDB 4 room flat ( ) HDB 5 room/Executive flat ( ) Condominium/Private apartment ( ) Landed property ( ) Others: ________ ( ) Prefer not to say |
| 10. Your Employment Status | ( ) Employed (full-time) ( ) Employed (part-time) ( ) Self-employed ( ) Unemployed ( ) Retired ( ) Homemaker ( ) Others: ________ ( ) Prefer not to say |
| 11. Your Average Monthly Household Income | ( ) Less than $2,000 ( ) $2,000 – $3,999 ( ) $4,000 – $5,999 ( ) $6,000 – $7,999 ( ) $8,000 and above ( ) Prefer not to say |
| 12. Your Marital Status | ( ) Single ( ) Married ( ) Widowed ( ) Divorced/Separated ( ) Prefer not to say |
